# Proteogenomic mapping of multimorbidity identifies C1R linking coronary artery disease and dementia

**DOI:** 10.64898/2026.07.14.26358022

**Authors:** Lin Li, Zhixin Tang, Zheng Zhong, Tingting Geng, Yanjun Guo, Yunfei Liao, Ayse Demirkan, Jack Bowden, Fiona Bragg, An Pan, Xufang Sun, Jiang Liu, Gang Liu, Jun Liu

## Abstract

Multimorbidity is highly prevalent in ageing populations, yet its shared molecular basis remains poorly defined, limiting the development of therapies that target multiple conditions. We systematically integrated measurements of 1,954 circulating proteins from 54,219 individuals in discovery and 35,559 in replication, focusing on ten common age-related diseases: coronary artery disease, chronic kidney disease, chronic obstructive pulmonary disease, dementia, heart failure, major depressive disorder, osteoarthritis, Parkinson’s disease, stroke, and type 2 diabetes. Coronary artery disease emerged as a central condition in the multimorbidity network, sharing circulating protein signatures with seven other diseases. Through genetic causal-inference analyses, we identified 40 circulating proteins with cross-disease relevance, of which four were further supported by colocalization of genetic variant associations. Among these, complement C1r, encoded by C1R, emerged as a key link between coronary artery disease and dementia, supported by independent colocalization evidence (PP.H4 = 0.86). Phenome-wide association analyses of C1R variants suggested that this signal was not driven by widespread unrelated genetic effects, but instead may reflect a more specific contribution to coronary artery disease–dementia pathogenesis. In vitro experiments further suggested that fibroblast-derived C1R promotes endothelial inflammation and neuronal apoptosis, providing mechanistic plausibility. Together, these findings position C1R as a biologically plausible and therapeutically relevant molecular link between coronary artery disease and dementia.

## Introduction

Multimorbidity, defined as the co-existence of two or more chronic conditions within an individual^1^, is highly prevalent in ageing populations and represents a major contributor to global morbidity and mortality^2,3^. Despite its growing clinical burden^4^, current medical practice remains largely organized around single-disease frameworks^5^, limiting the ability to address shared biological mechanisms across co-occurring conditions^3,6^. Identifying molecular pathways that operate across multiple conditions is therefore essential for developing more integrated and effective treatment strategies. The clinical co-occurrence of coronary artery disease (CAD) and dementia exemplifies this challenge. These two age-related conditions frequently coexist, and each independently increases the risk of the other, yet the molecular basis of their comorbidity remains poorly understood^7–10^.

Large-scale plasma proteomics offers a powerful approach to systematically identify shared molecular signals across diseases, as over 95% of approved drugs target proteins^11^. When combined with human genetic data, this approach becomes especially informative. Protein quantitative trait loci (pQTLs) can serve as strong instrumental variables for a causal inference approach, enabling assessment of whether genetically predicted circulating protein levels are associated with disease risk. This framework provides an opportunity to prioritize circulating proteins that may act across multiple disease pathways and serve as potential therapeutic targets for multimorbidity^11–13^.

Here, we performed a systematic, proteome-wide investigation across 1,954 plasma proteins from 54,219 individuals and 35,559 in replication, focusing on ten common age-related diseases which collectively account for a substantial proportion of the global disease burden^14^. Unlike single-disease proteomic studies, our approach maps shared protein-disease associations across multiple conditions and places functional validation at the center of target prioritization. By combining cross-disease proteomic evidence with genetic support and in vitro experiments, we identify targets with both population-level relevance and mechanistic plausibility. This framework systematically mapped shared molecular mediators across chronic diseases and nominated C1R as a candidate linking coronary artery disease and dementia (**Figure 1**)

**Figure 1.**
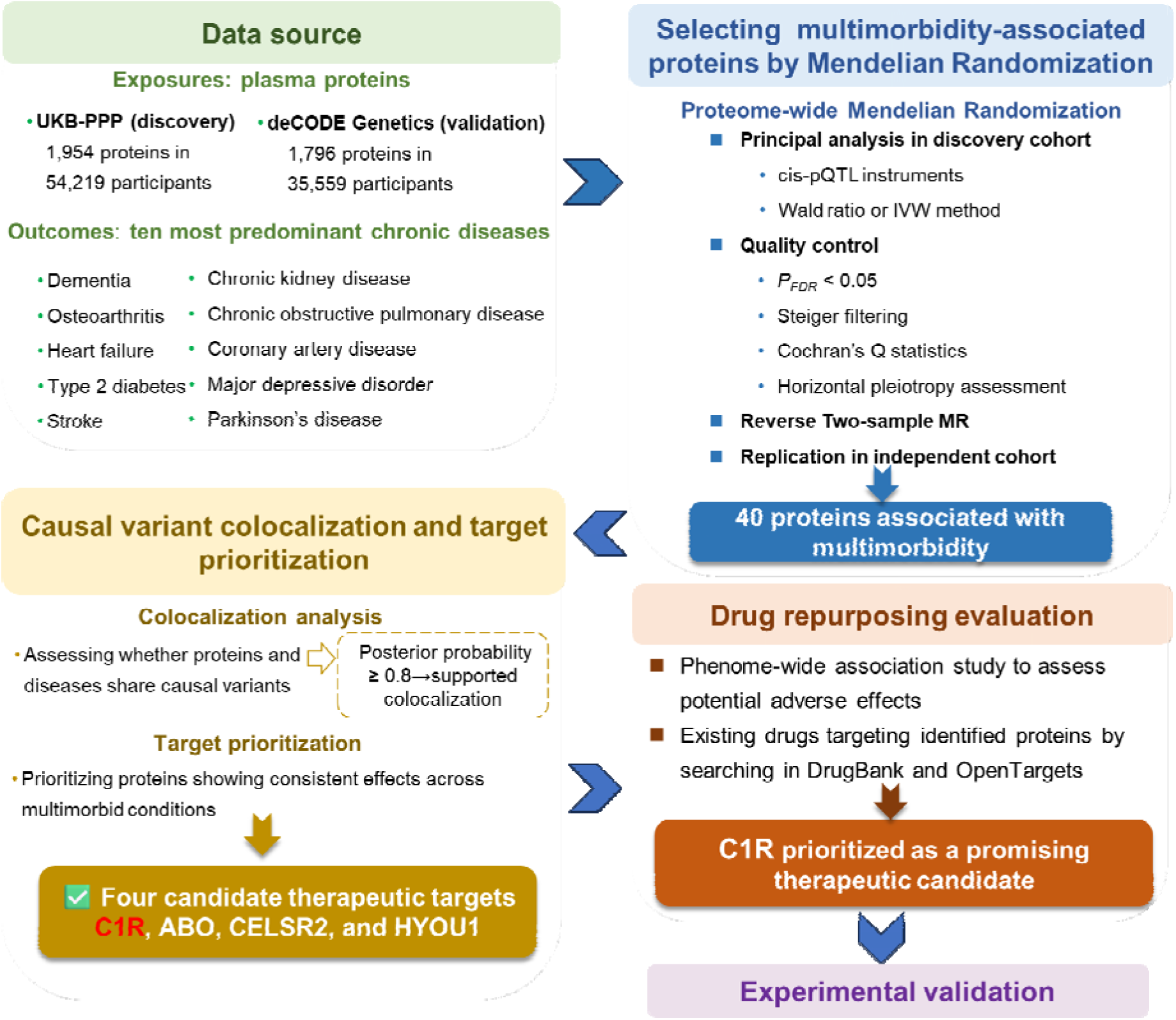
Overview of the study design. Abbreviation: FDR, False Discovery Rate; IVW, inverse variance-weighted; pQTL, protein quantitative trait loci

## Results

### A network of shared proteins links ten common chronic diseases

To systematically identify circulating proteins that may act across multiple chronic diseases, we performed two-sample Mendelian Randomization (MR) using cis-pQTLs for 1,954 plasma proteins (discovery cohort: UK Biobank, N=54,219)^15^ against ten common diseases: coronary artery disease (CAD), chronic kidney disease (CKD), chronic obstructive pulmonary disease (COPD), dementia, heart failure (HF), major depressive disorder (MDD), osteoarthritis (OA), Parkinson’s disease (PD), stroke, and type 2 diabetes (T2D) (**Table 1**)^16–26^. After multiple testing correction (*P_FDR_* < 0.05), genetically predicted levels of 224 proteins were significantly associated with at least one disease, yielding 274 protein–disease associations (**Supplementary Table 1**).

**Table 1.** Data sources for the 10 Chronic diseases and Proteins’ summary table.

|  | Sources | PubMed ID | Year | Ancestry | Cases/ controls |
| --- | --- | --- | --- | --- | --- |
| **Proteins <sup>a</sup>** |  |  |  |  |  |
| Plasma protein <sup>15, 29</sup> | UKB-PPP | 37794186 | 2022 | European | 54,219 |
|  | deCODE: PRJEB15197 | 34857953 | 2021 | European | 35,559 |
| **Chronic disease** |  |  |  |  |  |
| Dementia <sup>16,17</sup> | GWAS Catalog: GCST90027158 | 35379992 | 2022 | European | 85,934/401,577 |
|  | European Genome-phenome Archive: EGAS00001003424 <sup>b</sup> | 31473137 | 2019 | European | 4,120/3,289 |
| T2D <sup>24</sup> | DIAGRAM Consortium | 35551307 | 2022 | Multiancestry | 180,834/1,159,055 |
| OA <sup>23</sup> | GWAS Catalog: GCST007093 | 30664745 | 2019 | European | 77,052/378,169 |
| CKD <sup>22</sup> | GWAS Catalog: GCST008064 | 31152163 | 2019 | European and unknown | 64,164/561,055 |
| COPD <sup>21</sup> | GWAS Catalog: GCST90018807 | 34594039 | 2021 | European and East Asian | 17,547/617,598 |
| CAD <sup>18</sup> | GWAS Catalog: GCST90132314 | 36474045 | 2022 | European and unknown | 181,522/984,168 |
| MDD <sup>20</sup> | The Psychiatric Genomic Consortium | 29700475 | 2018 | Multiancestry | 67,162/454,450 |
| HF<sup>19</sup> | GWAS Catalog: GCST009541 | 31919418 | 2020 | European | 47,309/930,014 |
| PD <sup>26</sup> | International Parkinson's Disease Genomic Consortium | 31701892 | 2019 | European | 56,300/1,400,000 |
| Stroke <sup>25</sup> | MEGASTROKE consortium | 29531354 | 2018 | Multiancestry | 67,162/454,450 |

Abbreviation: OA, osteoarthritis; CKD, chronic kidney disease; COPD, chronic obstructive pulmonary disease; CAD, coronary artery disease; MDD, major depressive disorder; HF, heart failure; PD, Parkinson's disease; T2D, type 2 diabetes
<sup>a</sup> Proteomic data sources have no control groups; therefore, the sample sizes were the sample sizes used in each study.
<sup>b</sup> For dementia, the main dataset that was used in the study from Bellenguez C et al. <sup>16</sup>, which excluded the region corresponding to the APOE locus in this study. Since APOE is a well-known protein related to dementia, we added a study from Moreno-Grau et al. <sup>17</sup>, with a smaller sample size but including APOE, to complement the MR analysis between APOE and dementia.

Among these, 44 proteins showed genetically predicted associations with two or more diseases, collectively accounting for 89 protein–disease associations, indicating that protein sharing across conditions was a systematic feature rather than a rare phenomenon. Plasma levels of LPA, for example, exhibited genetically predicted positive associations with both CAD and HF, consistent with its well-established role in lipid metabolism and atherothrombotic cardiovascular disease^27,28^, providing a canonical example of pleiotropic protein–disease associations.

Reverse MR and MR-Steiger analyses supported the directionality of 88 of the 89 associations, leading to the exclusion of BTN3A2 (**Supplementary Table 2**). The remaining 43 proteins were subsequently evaluated in an independent Icelandic cohort (N=35,559, SomaLogic platform)^29^. Of these, 13 proteins were available for validation. Among them, ITIH3, MST1, and SF3B4 showed likely false-positive associations, as their MR results for at least one disease displayed inconsistent effects (*P_FDR_*<0.05 and associated in opposite direction) compared with those in the discovery cohort, leading to their exclusion for further analysis (**Supplementary Table 3**). Notably, five (C1R, C1S, NELL1, PRSS8, and SCARF2) showed consistent effect directions and statistical significance across their corresponding comorbid conditions using the same cis-pQTLs as in the discovery cohort (*P_FDR_* <0.05). For these five replicated proteins, we conservatively used an alternative set of cis-pQTL instruments derived from the Icelandic cohort GWAS summary statistics^29^ to evaluate the robustness of the replication. Among them, only C1R retained consistent positive associations with both CAD and dementia after FDR correction (*P_FDR_*< 0.05), indicating robustness to instrument selection (**Supplementary Table 4**).

### Coronary artery disease shows the most connections of protein-mediated multimorbidity

To understand whether protein sharing is organized around specific diseases, we constructed a network of the 40 validated proteins and their associated diseases. This network revealed CAD as the most highly connected node: 23 of the 40 proteins (57.5%) linked CAD to seven other conditions, including OA, T2D, dementia, HF, stroke, MDD, and CKD (**Figure 2A**). Pathway enrichment analysis of these CAD-connected proteins implicated the complement and coagulation cascades, suggesting that shared inflammatory and vascular mechanisms may underlie multimorbidity centered on CAD (**Figure 2B-C**).

**Figure 2.**
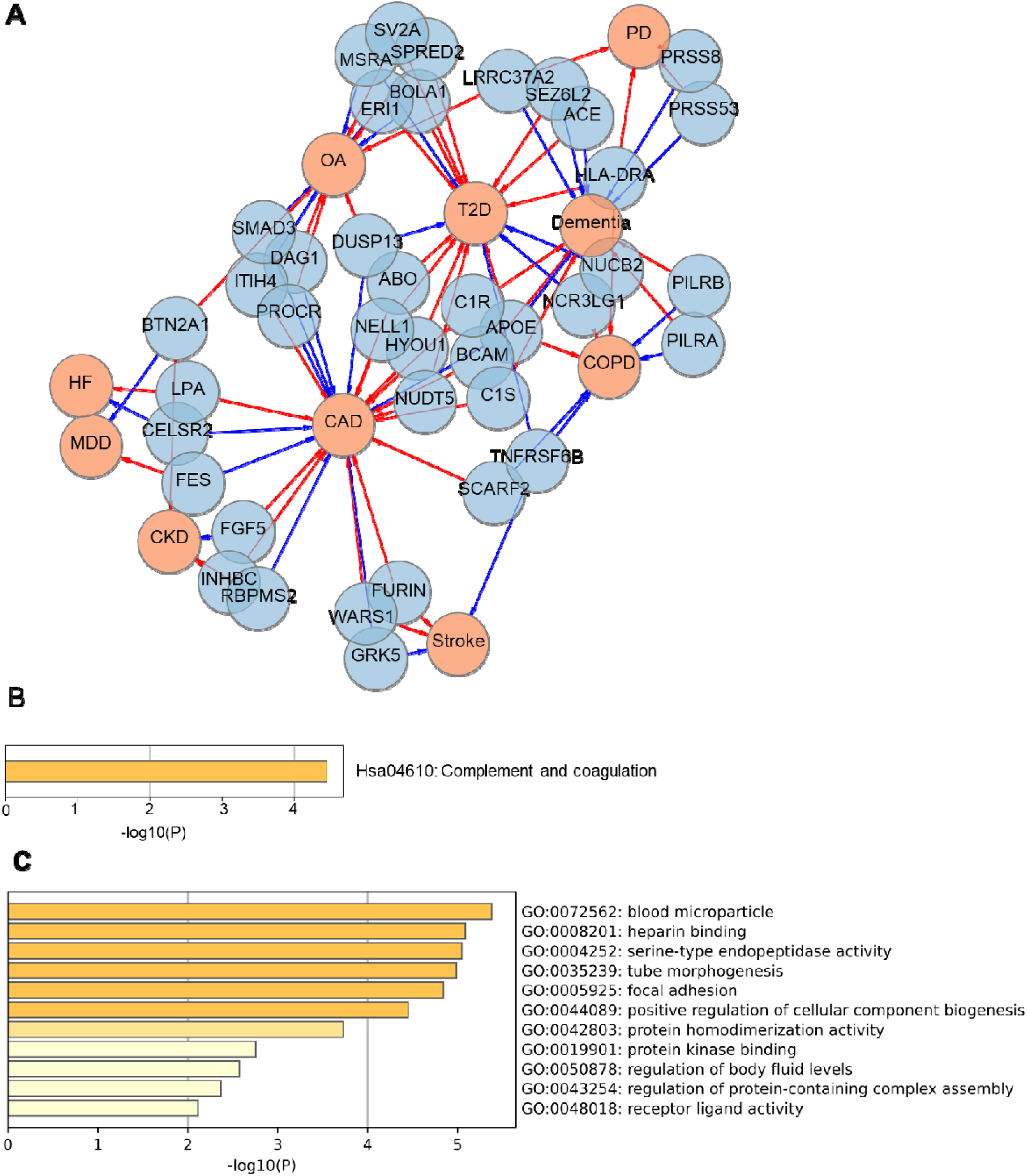
Contribution of CAD to multimorbidity-associated proteins and their functional enrichment. (A) Network of significant protein–multimorbidity associations, comprising 40 unique proteins and 87 protein–disease pairs. (B)-(C) Pathway enrichment analysis and Gene Ontology (GO) enrichment analysis of CAD-shared proteins. Abbreviation: OA, osteoarthritis; CKD, chronic kidney disease; COPD, chronic obstructive pulmonary disease; CAD, coronary artery disease; MDD, major depressive disorder; HF, failure; PD, Parkinson’s disease; T2D, type 2 diabetes.

### Genetic colocalization prioritizes four shared signals

To assess whether the identified protein–disease pairs share genetic determinants, we performed colocalization analysis. Of the 40 proteins, nine showed strong evidence of colocalization (posterior probability of H4 [PPH4] > 0.8)^30^. Among these, four had MR effect directions consistent across their associated diseases—C1R (CAD-dementia), ABO (CAD-T2D), HYOU1 (CAD-T2D), and CELSR2 (CAD-HF)—representing prioritized candidates for further evaluation (**Supplementary Table 5**).

C1R colocalized with both CAD and dementia (PPH4 = 0.86, driven by rs12146727 and rs10849546) and showed directionally consistent associations with increased risk of both diseases (**Figure 3A**). ABO, HYOU1, and CELSR2 also showed colocalization evidence (PPH4 = 0.99, 0.86, and 0.94, respectively) (**Figure 3B-D**). Two additional proteins (BTN2A1 and FGF5) colocalized but exhibited opposite directional effects across diseases, suggesting potential adverse effects if targeted (**Supplementary Figure 1**). Tissue expression profiling of the four prioritized candidates showed that C1R was expressed in both brain and heart tissues relevant to dementia and CAD, respectively—consistent with its potential dual role (**Supplementary Figure 2**)^31,32^.

**Figure 3.**
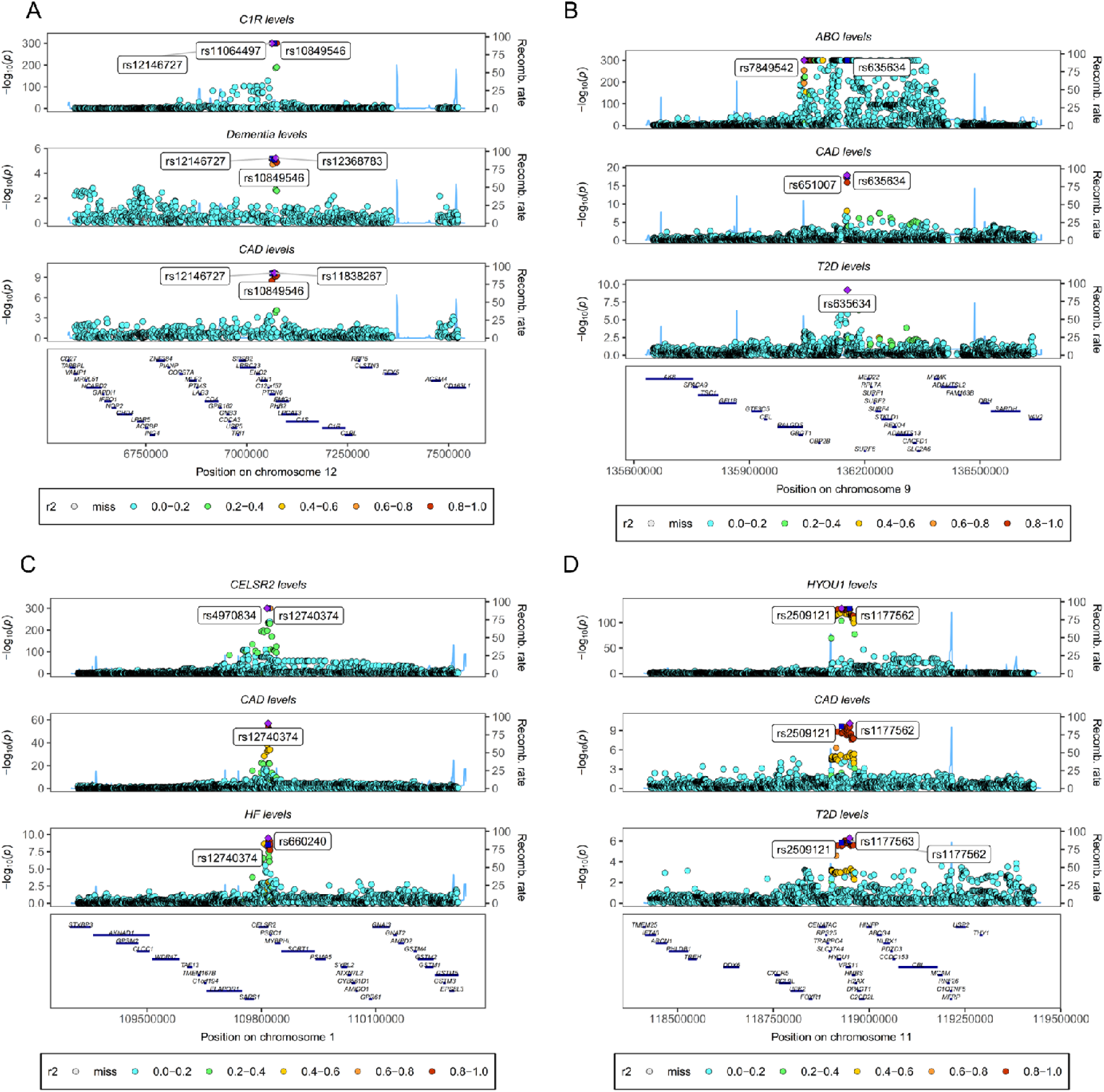
Colocalization between pQTL of identified protein targets and multimorbidity GWAS with high or moderate support of colocalization evidence. **(A)** Colocalization of the ABO pQTL signal in CAD and the T2D GWAS signal at the ABO locus. **(B)** Colocalization of C1R pQTL signal in CAD and dementia GWAS signal at the C1R locus. Colocalization of CELSR2 pQTL signal in CAD and HF GWAS signal at the CELSR2 locus. Colocalization of HYOU1 pQTL signal in CAD and T2D GWAS signal at the HYOU1 locus.

### Translational potential and PheWAS identify C1R as the lead candidate

We next evaluated the translational potential of the four candidates (C1R, ABO, HYOU1, and CELSR2) by systematic interrogation of DrugBank and OpenTargets databases^33,34^, combined with phenome-wide association studies (PheWAS) to assess on- and off-target effects (**Table 2 and Supplementary Tables 6-7**)^35^.

**Table 2.** Translational context of protein candidates with confirmed pharmacological actions.

| Gene | Drug name | Druggable Group | Type | Pharmacological action?/Actions | Current Indications |
| --- | --- | --- | --- | --- | --- |
| C1R | Conestat alfa | Approved | Biotech | YES/Inhibitor | Treat acute attacks of hereditary angioedema due to C1 esterase inhibitor deficiency in adults. |
| C1R | Human C1-esterase inhibitor | Approved | Biotech | YES/Inhibitor | Prevent angioedema attacks associated with hereditary angioedema. |
| C1R | Palivizumab | Approved | Biotech | YES/Activator | Prevent serious sequelae caused by respiratory syncytial virus infection in pediatric patients. |

<sup>a</sup> Data is extracted from the DrugBank (accessed in April 2025) and OpenTargets (accessed in June 2025).

C1R emerged as the most compelling target among the four candidates. Two clinically approved inhibitors—*conestat alfa* and *human C1-esterase inhibitor*—are currently used for hereditary angioedema^36,37^. Moreover, PheWAS revealed no significant associations of the C1R cis-pQTL allele with other diseases (**Figure 4**). In contrast, ABO showed pleiotropic associations with 63 traits, including protective effects against hypertension and pancreatic cancer, which would raise caution for inhibition. Multiple small-molecule drugs targeting ABO are in development, but their pharmacological actions and therapeutic indications remain unspecified. CELSR2 and HYOU1 currently lack documented therapeutic agents targeting their disease-associated functions, highlighting untapped opportunities for drug discovery. Consistent with the genetic findings, observational analyses in the UK biobank showed a positive association between circulating C1R levels and incident CAD after multivariable adjustment, whereas no significant association was observed for dementia (**Supplementary Figure 3**). Based on the integrated genetic evidence—genetic association and replication, colocalization, translational potential, and PheWAS—C1R was prioritized for experimental validation.

**Figure 4.**
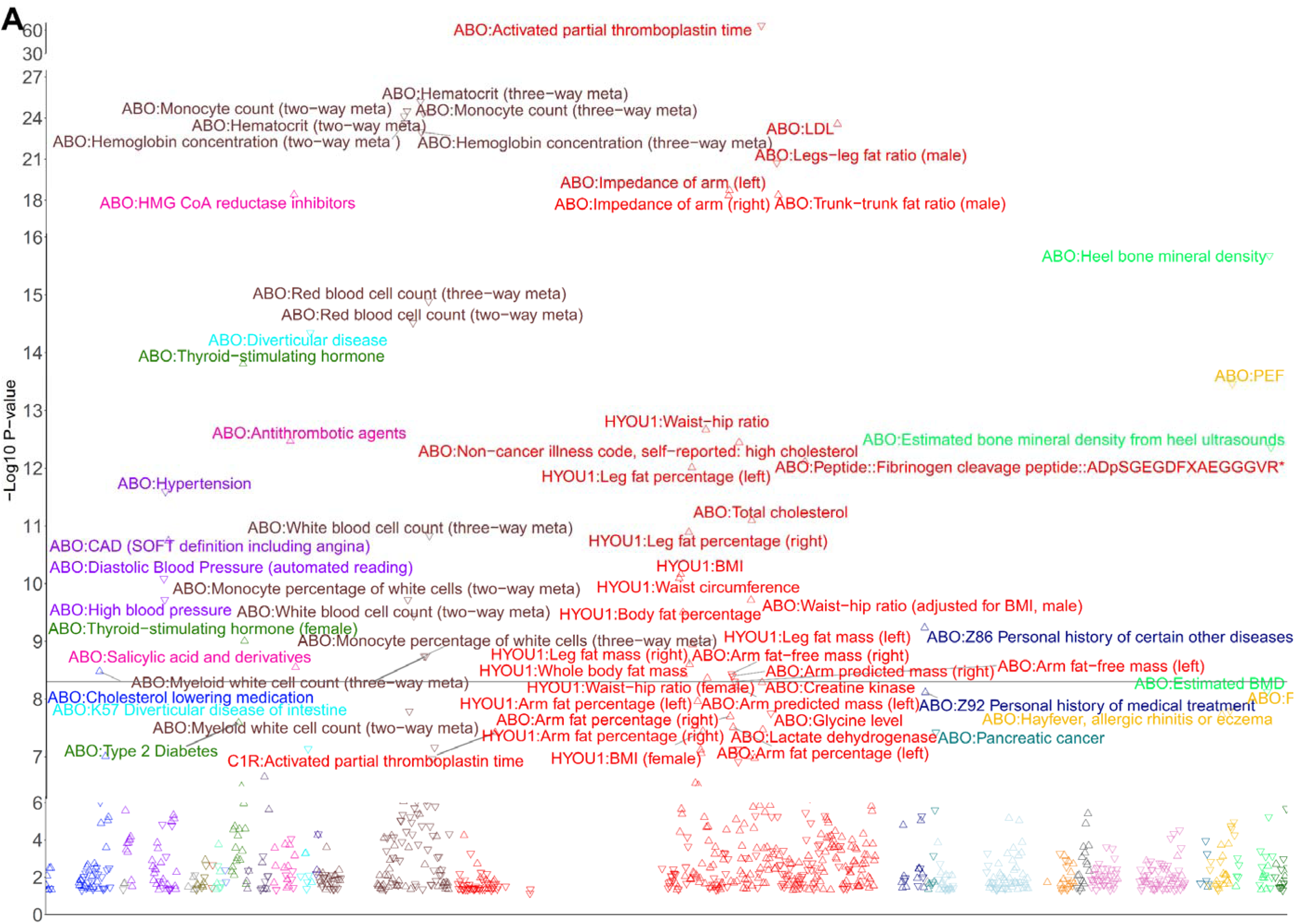
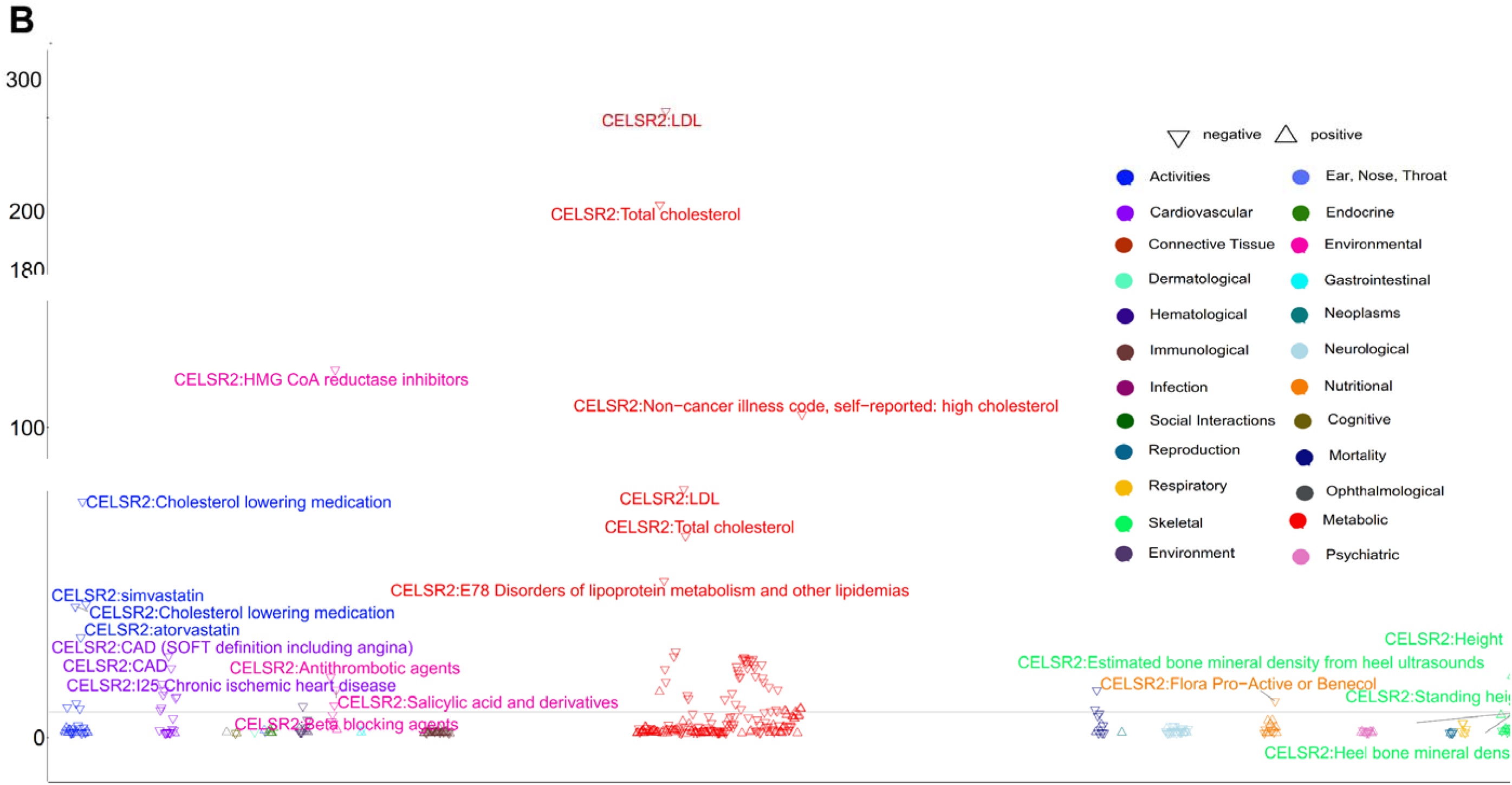
PheWAS plot for identified protein targets. (A) displayed the associations of top cis-pQTL allele of ABO (rs505922-C), C1R(rs10849546 and HYOU1(rs2509121-T) with phenotypes. (B) showed the associations of the top cis-pQTL allele of CELSR2 (rs12740374-T) with phenoty Triangles reflect associations of risk alleles of identified protein targets and traits, with triangles up representing positive associations and d representing negative associations. Triangle colors represent the categories to which traits belong.

### Single-cell and in vitro experiments support a paracrine role for fibroblast-derived C1R

To explore the cellular context of C1R in CAD and dementia, we analyzed single-cell transcriptomic data from human Alzheimer’s disease brain and atherosclerotic arteries^38,39^. Unexpectedly, C1R was predominantly expressed in fibroblasts in both tissues, whereas the upstream complement component C1QA was enriched in microglia and macrophages (**Figure 5 and Supplementary Figure 4**). This distinct cellular partitioning, together with MR analyses showing no genetic association between C1Q and either CAD or dementia, suggested that C1R may function independently of the canonical C1Q-initiated cascade.

**Figure 5.**
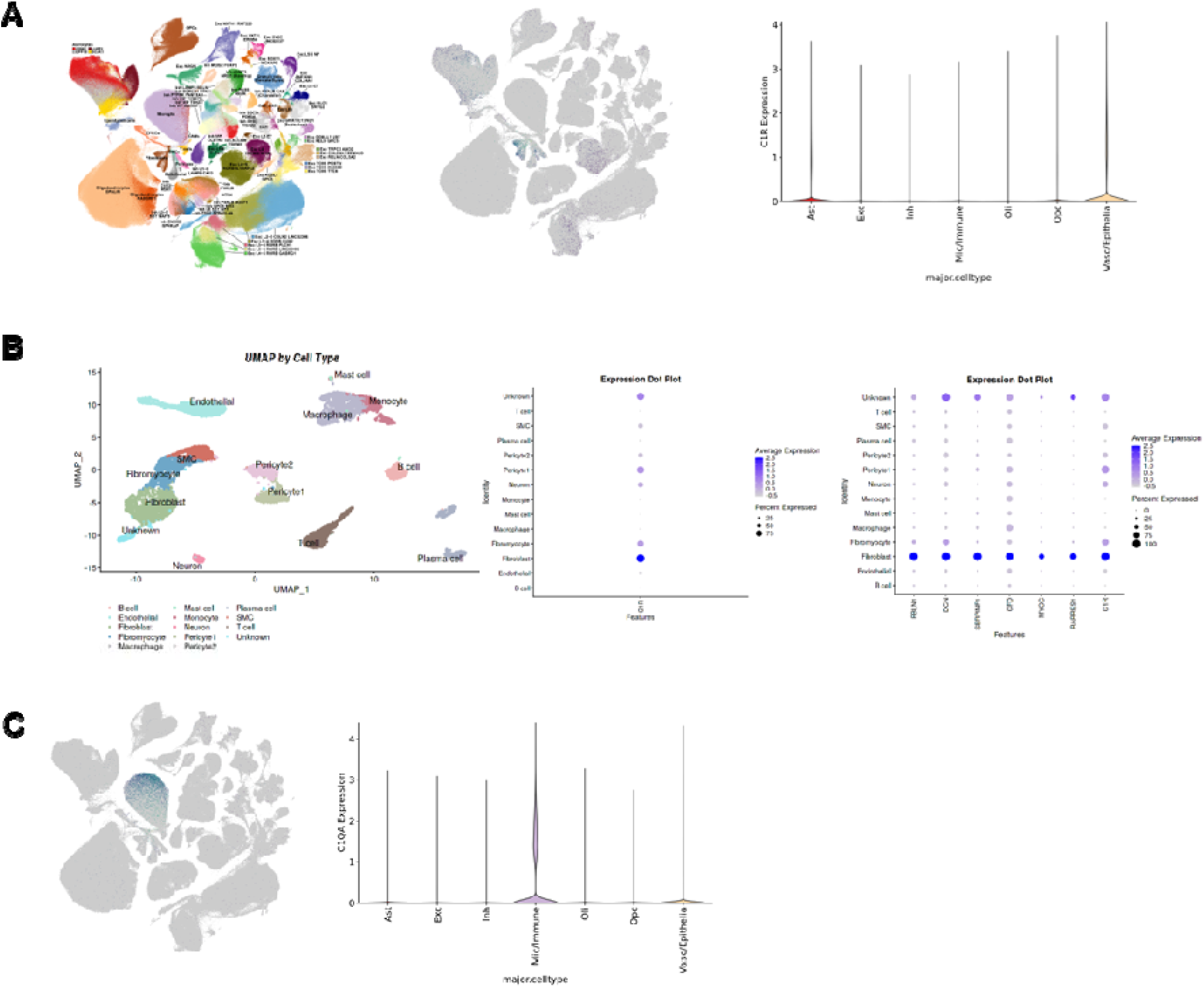
Expression of C1R in aging and aging-related diseases (A) UMAP and violin plots showing the cellular distribution of C1R in brain tissues from patients with Alzheimer’s disease using publicly available single-cell RNA sequencing data (http://compbio2.mit.edu/ad_multiregion/). (B) UMAP and violin plots illustrating the distribution of C1R in atherosclerotic vascular plaques using publicly available single-cell RNA sequencing data (https://plaqviewv2.pods.uvarc.io/). (C) UMAP and violin plots showing the cellular distribution of C1QA in brain tissues. Ast, astrocytes; Exc, excitatory neurons; Inh, inhibitory neurons; Mic/Immune, microglia/immune cells; Oli, oligodendrocytes; Opc, oligodendrocyte precursor cells; Vasc/Epithelia, vascular/epithelial cells.

In human vascular adventitial fibroblasts (HVAFs), inflammatory or oxidative stress did not alter intracellular C1R levels but markedly enhanced its secretion into the extracellular environment (**Supplementary Figure 5**). Exogenous recombinant C1R potentiated ox-LDL-induced upregulation of endothelial adhesion molecules (VCAM-1 and E-selectin) in human umbilical vein endothelial cells and exacerbated Aβ25–35-induced apoptosis in SH-SY5Y neuronal cells (**Figure 6A-C**). To test whether fibroblast-derived C1R acts in a paracrine manner, we established a Transwell co-culture system. Silencing C1R in HVAFs significantly reduced endothelial activation under ox-LDL stimulation and attenuated neuronal apoptosis under Aβ25–35 exposure (**Figure 6D-F**). These findings suggest that fibroblast-derived C1R acts as a paracrine mediator that amplifies tissue injury in both vascular and neurodegenerative contexts.

**Figure 6.**
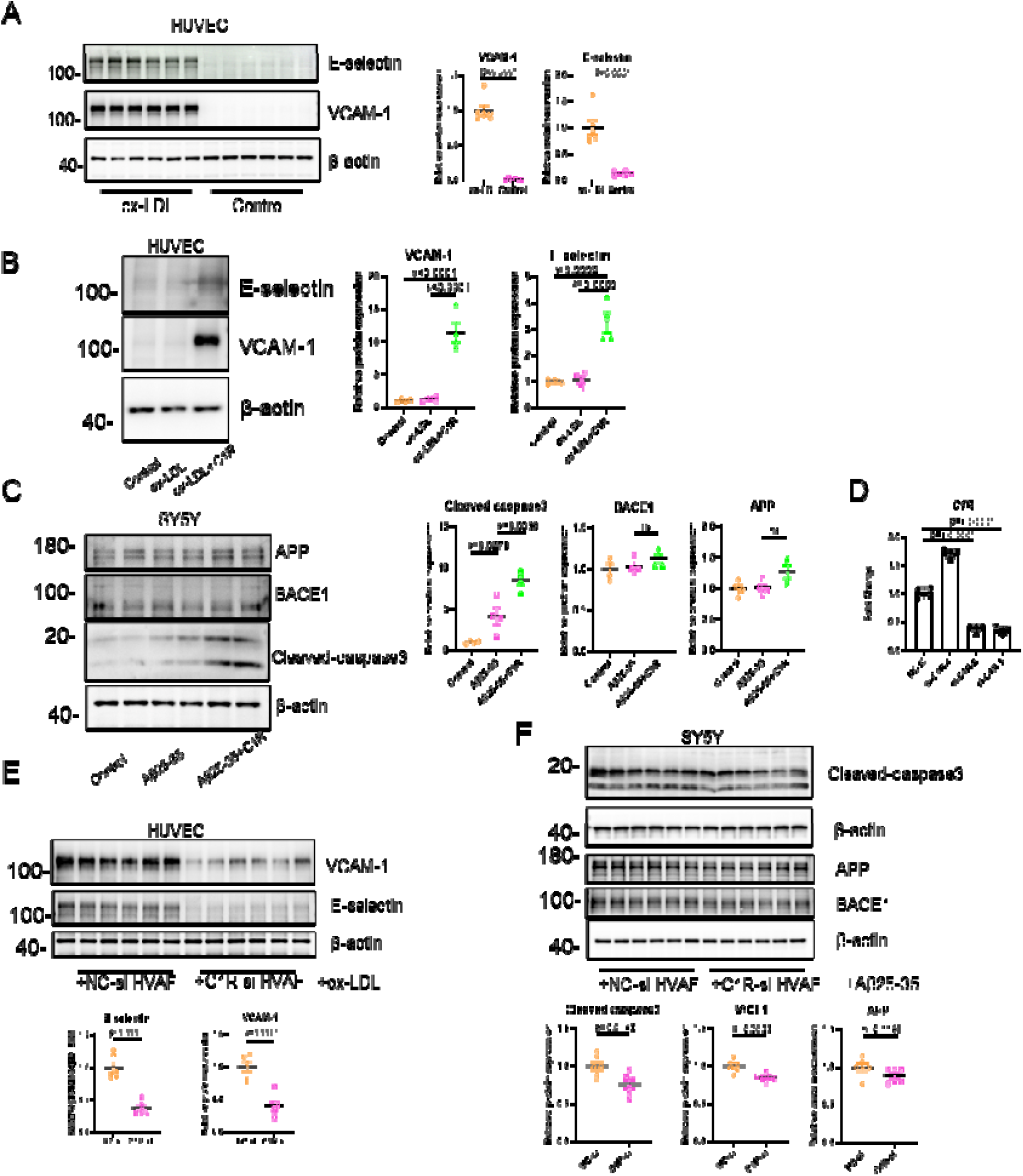
C1R exacerbates vascular endothelial inflammation and neuronal injury (A) Western blot quantification of E-selectin and VCAM-1 protein expression in HUVECs following ox-LDL stimulation (n = 6 per group). (B) Western blot quantification of E-selectin and VCAM-1 expression in HUVECs after treatment with recombinant C1R (n = 4 per group). (C) Western blot quantification of APP, BACE1, and cleaved caspase-3 protein expression in SH-SY5Y cells following treatment with recombinant C1R (n = 4 per group). (D) Validation of C1R knockdown efficiency in HVAFs by RT-qPCR (n = 4 per group). (E) Western blot quantification of E-selectin and VCAM-1 expression in HUVECs co-cultured with HVAFs transfected with NC-si or C1R-si under ox-LDL stimulation (n = 6 per group). (F) Western blot quantification of APP, BACE1, and cleaved caspase-3 expression in SH-SY5Y cells co-cultured with HVAFs transfected with NC-si or C1R-si under Aβ25–35 stimulation (n = 6 per group). Data are presented as mean ± SEM. Statistical significance between the two groups was analyzed using an unpaired t-test. Statistical comparisons among multiple groups were performed using one-way ANOVA with Bonferroni’s multiple comparisons test.

## Discussion

The coexistence of coronary artery disease and dementia in aging populations represents one of the most pressing clinical challenges of multimorbidity, yet no molecular link has been systematically established. Here, through a proteome-wide genetic study across ten common chronic diseases, we identify C1R as a shared circulating protein that genetically and experimentally connects CAD to dementia, and for which clinically approved inhibitors already exist.

The finding that CAD shows the most connections in the protein-mediated multimorbidity network carries both biological and clinical implications. More than half of the shared proteins connecting CAD to other conditions—ranging from dementia to OA and CKD—are enriched in the complement and coagulation cascades, key regulators of inflammation and thrombosis that are central to atherosclerosis^40–44^. This enrichment suggests that vascular injury and low-grade systemic inflammation, rather than organ-specific pathology, may represent a common soil on which multimorbidity develops. Clinically, this network topology predicts that individuals with CAD are at risk not merely for a single secondary condition but for a cluster of diverse diseases linked through shared inflammatory mediators. This pattern aligns with epidemiological evidence that CAD often coexists with multiple diseases across various organ systems in aging populations^7–10^. Consequently, protein-targeted interventions in CAD patients might yield unexpected benefits across multiple organ systems, a hypothesis that prospective trials with proteomic stratification could test.

Among the 40 shared proteins, C1R stood out for its convergent genetic evidence linking higher risk of both CAD and dementia. While previous studies separately implicated C1R protein in dementia and the C1R locus in CAD^45,46^, our study provides the first evidence—to our knowledge—that circulating C1R may represent a common molecular factor underlying their comorbidity. C1R is a serine protease that initiates the classical complement pathway through formation of the C1 complex with C1Q and C1S^47^, and aberrant activation of this pathway has been implicated in chronic inflammatory processes across multiple diseases^48,49^. Our findings raise the possibility that C1R may act as a molecular bridge linking vascular and neurodegenerative pathology through complement-mediated inflammatory signaling.

Given the strong age-dependent co-occurrence of CAD and dementia^50^, we investigated the cellular source of C1R in disease-relevant tissues. Using single-cell transcriptomic data from Alzheimer’s disease brain and atherosclerotic arteries, we observed that C1R was predominantly expressed in fibroblasts, whereas the upstream complement component C1Q was enriched in microglia and macrophages^38,39,51^. This distinct cellular partitioning suggests a potential division of labor within the classical complement pathway: immune cells may initiate complement signaling while fibroblasts provide a local source of C1R. This finding raises the possibility that fibroblast-derived C1R contributes to pathological processes shared between vascular and neurodegenerative diseases, potentially independent of the canonical C1Q-initiated cascade—a notion supported by our MR analysis showing no genetic association between C1Q and either CAD or dementia. Although genetically predicted C1R levels showed consistent associations with CAD and dementia across multiple genetic frameworks and experimental validation, circulating C1R protein levels were not significantly associated with dementia in observational analyses. This discrepancy may reflect differences in temporal scale, as genetic instruments capture lifelong exposure, whereas circulating protein measurements reflect short-term variation that is susceptible to unmeasured confounding and reverse causation^29,52^. These findings suggest that C1R may influence disease susceptibility primarily through long-term regulation of complement activity rather than reflecting acute circulating protein levels.

Our in vitro experiments further support a paracrine role for fibroblast-derived C1R. Under inflammatory or oxidative stress, fibroblasts increased C1R secretion, and exogenous C1R potentiated endothelial activation and neuronal apoptosis. Conversely, silencing C1R in fibroblasts attenuated both responses in co-culture systems. These findings suggest a model in which fibroblast-derived C1R acts locally to amplify tissue injury in both vascular and neurodegenerative contexts, providing a potential mechanistic link between CAD and dementia at the level of the tissue microenvironment.

From a therapeutic perspective, C1R offers several advantages. Clinically approved inhibitors targeting the classical complement pathway—conestat alfa and human C1 esterase inhibitor—are already used for hereditary angioedema^53,54^, providing proof-of-concept for pharmacological modulation. However, these agents have not been approved for CAD or dementia, and their efficacy and long-term safety in these conditions remain to be established. Our PheWAS analysis revealed no significant associations of the C1R cis-pQTL allele with other diseases. However, because C1R is an integral component of innate immunity^47^, prolonged systemic suppression could theoretically increase infection susceptibility^55^. Encouragingly, our PheWAS did not identify strong associations with infection-related phenotypes, suggesting that moderate modulation may be tolerated. Given the substantial comorbidity burden between CAD and dementia^56^, carefully titrated complement modulation could offer a shared therapeutic avenue for both conditions, warranting further evaluation in preclinical models.

Beyond C1R, our systematic screening identified four additional proteins—ABO, HYOU1, and CELSR2—with robust genetic evidence supporting their involvement in multiple cardiometabolic diseases. Notably, these candidates fell into two directional patterns: higher ABO and HYOU1 levels were associated with increased disease risk, whereas higher CELSR2 levels showed protective associations. This bidirectional pattern is biologically plausible—ABO and HYOU1 link to pro-inflammatory and stress-response pathways^57–60^, while CELSR2 has been implicated in lipid regulation and cardiac stress adaptation^61–63^. These findings suggest that multimorbidity may arise not only from shared risk factors but also from the breakdown of shared protective mechanisms.

Several limitations should be acknowledged. First, our analyses focused on circulating proteins; tissue-specific proteomic data may provide additional insights. Second, while cis-pQTLs minimize horizontal pleiotropy, they cannot be completely excluded, and platform-related discrepancies between discovery and replication cohorts may exist^64^. Third, our multimorbidity framework is inherently shaped by the availability and power of GWAS data for the ten diseases studied, potentially overrepresenting well-powered conditions. Fourth, while our in vitro experiments demonstrate that fibroblast-derived C1R amplifies endothelial and neuronal injury, the precise molecular mechanisms remain to be elucidated. In vivo loss-of-function studies using C1R knockout mice or pharmacological inhibition in relevant disease models will be essential to establish causality and validate therapeutic potential. Fifth, PheWAS based on a cis-pQTL cannot fully recapitulate pharmacological inhibition, and the favorable short-term profile of C1R inhibitors in hereditary angioedema may not extend to chronic suppression in older multimorbid adults.

In summary, integrating large-scale plasma proteomics with genetic causal inference enabled systematic mapping of shared proteomic signals across ten major chronic diseases. Among the identified candidates, C1R emerged as a prioritized mediator linking CAD and dementia, supported by convergent genetic evidence. Notably, clinically approved C1R inhibitors already exist for hereditary angioedema, raising the possibility of therapeutic repurposing for CAD-dementia multimorbidity. While in vivo studies are needed to establish causality, these findings highlight complement signaling as a shared pathway across cardiometabolic and neurodegenerative diseases and illustrate the potential of proteomics-guided causal inference to uncover therapeutic opportunities for age-related multimorbidity.

## Methods

### Proteomic data source

In this study, we utilized proteomic data from two independent, non-overlapping studies by Sun et al. and Ferkingstad et al.^15,29^, as discovery and validation datasets, respectively. The discovery dataset comprised plasma proteomic profiles from 54,219 participants in the UK Biobank, quantified using the Olink Explore platform^15^. Sun et al. mapped pQTLs for 2,923 proteins, identifying 14,287 primary genetic associations. Among these, 1,954 proteins were associated with 1,955 cis-pQTLs within a 1Mb range at a significance threshold of *P* < 1.7×10^-11^. For validation, we used data from Ferkingstad et al.’s study involving 35,559 Icelandic individuals, where plasma proteins were measured using 4,907 aptamers on the SomaScan platform^29^. This study identified 28,191 pQTL associations, including both sentinel and secondary associations (*P* < 1.8×10^-9^), with 1,796 proteins linked to 7,572 cis-pQTLs within a 1Mb range.

### Disease data sources

The ten most predominant chronic diseases are dementia, OA, CKD, COPD, CAD, MDD, HF, PD, stroke, and T2D^15–26,29^. The summary-level statistics of genetic associations with these ten diseases were extracted from the largest available existing GWAS, and the details are shown in **Table 1**. For dementia, the main dataset that was used in the study from Bellenguez C et al. ^16^, which excluded the region corresponding to the APOE locus in this study. Since APOE is a well-known protein related to dementia, we added a study from Moreno-Grau et al. ^17^, with a smaller sample size but including APOE, to complement the MR analysis between APOE and dementia.

### MR analysis

To assess the causal effect of circulating proteins on ten specified diseases, we utilized a two-sample MR approach. In the discovery dataset, genetic instruments were selected from published cis-pQTLs identified in the previous large-scale proteome-wide GWAS study^15^. For each protein-disease pair, we applied the Wald ratio method when a single SNP was used as the instrumental variable (IV), and the IVW method when multiple independent SNPs were available^65^. To account for multiple testing, FDR correction was applied to obtain adjusted *P* values for each protein-disease association, with FDR-adjusted *P* < 0.05 considered statistically significant. To ensure the robustness of causal estimates, we performed sensitivity analyses only when multiple SNPs were available, including diagnostic tests for pleiotropy (MR–Egger intercept test), directionality (the Steiger directionality test), and heterogeneity (Cochran’s Q statistics). Associations with *P* values > 0.05 in all three tests were considered robust and free of substantial pleiotropy, reverse causation, or heterogeneity. Using a conservative threshold, we excluded any association that failed one or more of these diagnostic tests (*P* < 0.05) from further analyses. Proteins that showed significant associations with two or more diseases (FDR-adjusted *P* < 0.05 for each, and passing all sensitivity analyses) were considered as shared causal factors across multimorbidity, and included in downstream analyses.

To further rule out reverse causation, we performed reverse MR analyses for proteins significantly associated with multimorbidity. The selection of SNPs used as IVs was based on the following criteria: SNPs meet the genome-wide significance threshold (*P* < 5 × 10^-8^), and linkage disequilibrium (LD) clumping was performed using a 10,000 kb window and r^2^ < 0.01 based on the European 1000 Genomes Project panel^66^. Reverse MR analyses and sensitivity assessments followed the same methodological framework as described above and were implemented using the utilizing TwoSampleMR R package^67^.

For the proteins identified in the discovery dataset (UK Biobank), protein–disease associations were further validated in the Iceland cohort if applicable. IVs were selected from previously published cis-pQTLs identified in the Icelandic cohort ^29^. The selection process was as follows: (1) If the IVs identified for a given protein in the Iceland cohort overlapped with those used for the same protein in the UK Biobank, the overlapping instruments were prioritized to ensure consistency with the discovery analysis; (2) If no overlapping IVs were available, we selected instruments based on the following criteria: first, cis-pQTLs that were present in both the protein and the corresponding comorbid disease were identified; then, the cis-pQTL showing the strongest association with the protein (i.e., the smallest p-value) was selected as the instrument for Mendelian randomization (MR) analysis; (3) If none of the above criteria were met, the protein was excluded from the validation analysis. For the initially replicated associations, sensitivity analyses were performed using the alternative cis-pQTL instruments derived from the Icelandic cohort GWAS summary statistics^29^.

### Functional and pathway enrichment analysis

Kyoto Encyclopedia of Genes and Genomes (KEGG) pathway analysis and Gene ontology enrichment analysis, including GO-BP, GO-CC, and GOMF, were performed for the identified proteins using the Metascape platform^68^.

### Multi-trait colocalization analysis

While MR largely mitigates bias from confounding, LD between SNPs may still introduce noncausal associations. To address this, multi-trait colocalization analysis using HyPrColoc method^30^ was performed for proteins that showed shared associations with multimorbidity. This approach estimates the probability that both the protein and associated diseases share a single causal variant, thereby assessing potential confounding due to LD. We analyzed all SNPs within 1 Mb of the lead cis-pQTL and cis-GWAS of corresponding diseases. Colocalization analysis was performed using the hyprcoloc R package^69^, and the results were visualized with the gassocplot2 R package^70^. A PP of more than 0.8 (the probability of a shared causal variant) was considered as the association with high support of colocalization evidence. Proteins with consistently genetically predicted associations across multimorbid conditions, supported by high or moderate colocalization probabilities, were prioritized as protein candidates for multimorbidity.

### Gene expression analyses

Gene expression profiles of the encoding genes of the identified protein candidates were examined using the default GENE2FUNC pipeline in FUMA, based on the GTEx v8 dataset^31,32^. Raw expression values were normalized by adding a pseudo-count of 1 and then log2-transformed to calculate average expression levels across 30 different tissues. Genes with log2-transformed expression values greater than 2.84, corresponding to an expression level approximately 6.5-fold higher than the median expression level across all genes, were considered to be highly expressed^32,71^.

### Phenome-wide association study

PheWAS is a method employed to investigate associations between SNPs or phenotypes and a broad spectrum of phenotypes across the entire phenome^72^, which is especially valuable for exploring potential adverse effects associated with drug targets^73^. We further studied the pleiotropic effect of the genetic determinants of the identified drug targets using PheWAS by data mining from previous publications in GWAS ATLAS ^35^, including a total of 4756 traits from 28 phenome categories. The *P* threshold of < 5 × 10^-8^ was considered statistically significant.

### Therapeutic potential of identified drug targets

To evaluate the therapeutic potential of identified proteins, we searched for identified proteins with the same causal direction as multiple diseases in DrugBank and OpenTargets^33,34^. For proteins identified in the drug database, information on drug names, molecule types, approved indications, and target outcomes in clinical trials was documented. Moreover, to assess the potential for drug targeting, we categorized these proteins into four groups: 1) Approved: proteins for which one or more drugs targeting them have been approved; 2) In clinical trials: proteins targeted by drugs currently under investigation in clinical trials; 3) Preclinical: proteins targeted by drugs in the preclinical development stage; 4) Protein candidate: proteins not identified in drug databases but considered potential targets for drug development.

### Cox proportional hazards regression analyses in the UK Biobank

Time-to-event analyses were conducted using Cox proportional hazards regression models to estimate hazard ratios (HRs) and 95% confidence intervals (CIs) for the associations of plasma C1R levels with incident CAD and dementia in the UK Biobank cohort, which was used as an independent validation population for the prioritized protein targets identified from genetic analyses. Follow-up time was calculated from baseline assessment to the date of outcome diagnosis, death, loss to follow-up, or end of follow-up, whichever occurred first. We constructed a series of hierarchical models with progressive adjustment for potential confounders. Model 1 adjusted for age (continuous), sex (male or female), self-reported race (White, Asian or Asian British, Black or Black British, or other), and study center (England, Wales, or Scotland). Model 2 further adjusted for educational attainment (college or university degree, intermediate qualifications, or low qualifications) and Townsend deprivation index (continuous). Model 3 additionally adjusted for body mass index (<25, 25–30, or ≥30 kg/m²), alcohol consumption (never, previous, or current), smoking status (never, former, or current), and physical activity (low, moderate, or high). Model 4 further adjusted for baseline history of hypertension, hyperlipidemia, and cancer (yes or no). Model 5 additionally adjusted for cardiovascular disease (yes or no), social isolation (yes or no), sleep quality (good or poor), depressive symptoms (yes or no), family history of dementia (yes or no), and APOE ε4 carrier status.

### In vitro experiments for C1R

Human umbilical vein endothelial cells (HUVECs; ZQ1099, RRID CVCL_F0BB) were maintained in Complete Medium (Zhong Qiao Xin Zhou Biotechnology). Human neuroblastoma SH-SY5Y cells (CRL-2266, ATCC) were cultured in DMEM (Gibco) with 10% fetal bovine serum (Cegrogen). Human vascular adventitial fibroblasts (HVAFs; CM-H200, Procell) were cultured in Complete Medium (CM-P200, Procell) and used at passages 3–5. All cells were maintained at 37°C with 5% CO_2_.

For co-culture, HVAFs were seeded in the upper chamber of Transwell inserts (0.4 μm pore size; Labselect), with HUVECs or SH-SY5Y cells in the lower chamber. HVAFs were transfected with siRNA using Lipofectamine MessengerMAX (Thermo Fisher Scientific). siRNA sequences are listed in **Supplementary Table 8**.

Protein samples were separated by SDS–PAGE and transferred to PVDF membranes (GenScript). Membranes were blocked with 5% non-fat milk in TBST, incubated with primary antibodies overnight at 4°C, then with HRP-conjugated secondary antibodies (1:10,000; Aspen). Bands were detected using enhanced chemiluminescence and quantified with ImageJ. For serum samples, albumin and immunoglobulin were removed using a depletion kit (P2295, Beyotime) prior to analysis. Primary antibodies used: anti-C1R (GTX104350), anti–E-selectin (20894-1-AP), anti–VCAM-1 (11444-1-AP), anti-BACE1 (12807-1-AP), anti-APP (25524-1-AP), anti–cleaved caspase-3 (9664), anti–β-actin (AC026), and anti-GAPDH (60004-1-Ig) (see **Supplementary Table 9** for sources and catalog numbers).

Total RNA was extracted using an RNA extraction kit (RC112, Vazyme) and reverse-transcribed using a reverse transcription kit (R223, Vazyme). qPCR was performed using SYBR Green Master Mix (Q711, Vazyme) with GAPDH as an internal control. Relative expression was calculated using the 2−ΔΔCt method. Primer sequences are listed in **Supplementary Table 10.**

For the cell viability assay, SH-SY5Y cells were seeded in 96-well plates (5 × 10³ cells/well) and treated with HFIP-pretreated Aβ25–35 (HY-P0128, MedChemExpress) for 48 h. Cell viability was assessed using Cell Counting Kit-8 (ABclonal) by measuring absorbance at 450 nm.

Data are presented as mean ± SEM. Normality was assessed using the Shapiro-Wilk test. Comparisons between two groups used a two-tailed Student’s t-test; multiple comparisons used one-way ANOVA with Bonferroni correction. A minimum of three independent biological replicates was performed. Analyses were conducted in GraphPad Prism (v9.0.0).

## Supporting information

uncropped gels

## Author contributions

J.L., G.L., J.L. (Jiang Liu), and X.S. are the guarantors of this work and, as such, have full access to all the data in the study and take responsibility for the integrity of the data and the accuracy of the data analysis. J.L. (Jiang Liu) conceived and designed the content on protein function and therapeutic potential. X.S. and J.L. (Jun Liu) conceived and designed the experimental work. J.L. (Jun Liu) and G.L. conceived and designed the remainder of the study. L.L., Z.T., and Z.Z. contributed equally as co-first authors. L.L. and Z.T. undertook the statistical analyses, interpreted the data, and wrote the first draft of the manuscript under the supervision of J.L. (Jun Liu), G.L., and J.L. (Jiang Liu). Z.Z. performed all experimental work and drafted the corresponding portion of the manuscript under the supervision of X.S. All authors (L.L., Z.T., Z.Z., T.G., Y.G., Y.L., A.D., J.B., F.B., A.P., X.S., J.L. [Jiang Liu], G.L., and J.L. [Jun Liu]) contributed to the interpretation of the results and critical revision of the manuscript for important intellectual content. All authors approved the final version of the manuscript.

## Acknowledgments

The authors acknowledge the important contributions of all publicly available data sets used in this report’s analyses.

## Data availability

Our pQTL summary data were acquired from previously published studies and can be found in the supplemental materials of these studies (https://www.nature.com/articles/s41586-023-06592-6, and https://www.nature.com/articles/s41588-021-00978-w). The GWAS data for diseases including dementia, Osteoarthritis, Chronic Kidney disease, Chronic obstructive pulmonary disease, Coronary artery disease, and heart failure, are publicly available in the GWAS Catalog under accession codes: GCST90027158, GCST007093, GCST008064, GCST90018807, GCST90132314, and GCST009541. The summary data of other diseases, including Type 2 diabetes, Major Depression disease, Parkinson’s disease, and stroke, were extracted from DIAGRAM Consortium [https://www.diagram-consortium.org/], The Psychiatric Genomic Consortium [https://pgc.unc.edu/], International Parkinson’s Disease Genomic Consortium [https://www.pdgenetics.org/], MEGASTROKE consortium [https://megastroke.org/]. Other data sources for dementia and CAD are publicly available in the European Genome-phenome under accession code: EGAS00001003424 and IEU OpenGWAS [https://opengwas.io/]. The Open Targets data are deposited in https://platform.opentargets.org/. The DrugBank data are deposited in https://go.drugbank.com/. The FUMA data are deposited in https://fuma.ctglab.nl/gene2func/. The observational analysis in this study has been conducted using the UK Biobank resource under application number 109546. UK Biobank data are available to all researchers for health-related research and public interest through registration on the UK Biobank (www.ukbiobank.ac.uk).

## Code Availability

The analytic code used in this study will be made available upon request.

## Ethics approval

The UK Biobank had ethics approval from the North West Multi-Centre Research Ethics Committee, the National Information Governance Board for Health and Social Care in England and Wales, and the Community Health Index Advisory Group in Scotland. Informed consent was obtained from all participants. The other part of the study is based on publicly available data, which had been approved by the corresponding ethical review committees.

