## Supplementary figures for "Proteogenomic mapping of multimorbidity identifies C1R linking coronary artery disease and dementia"

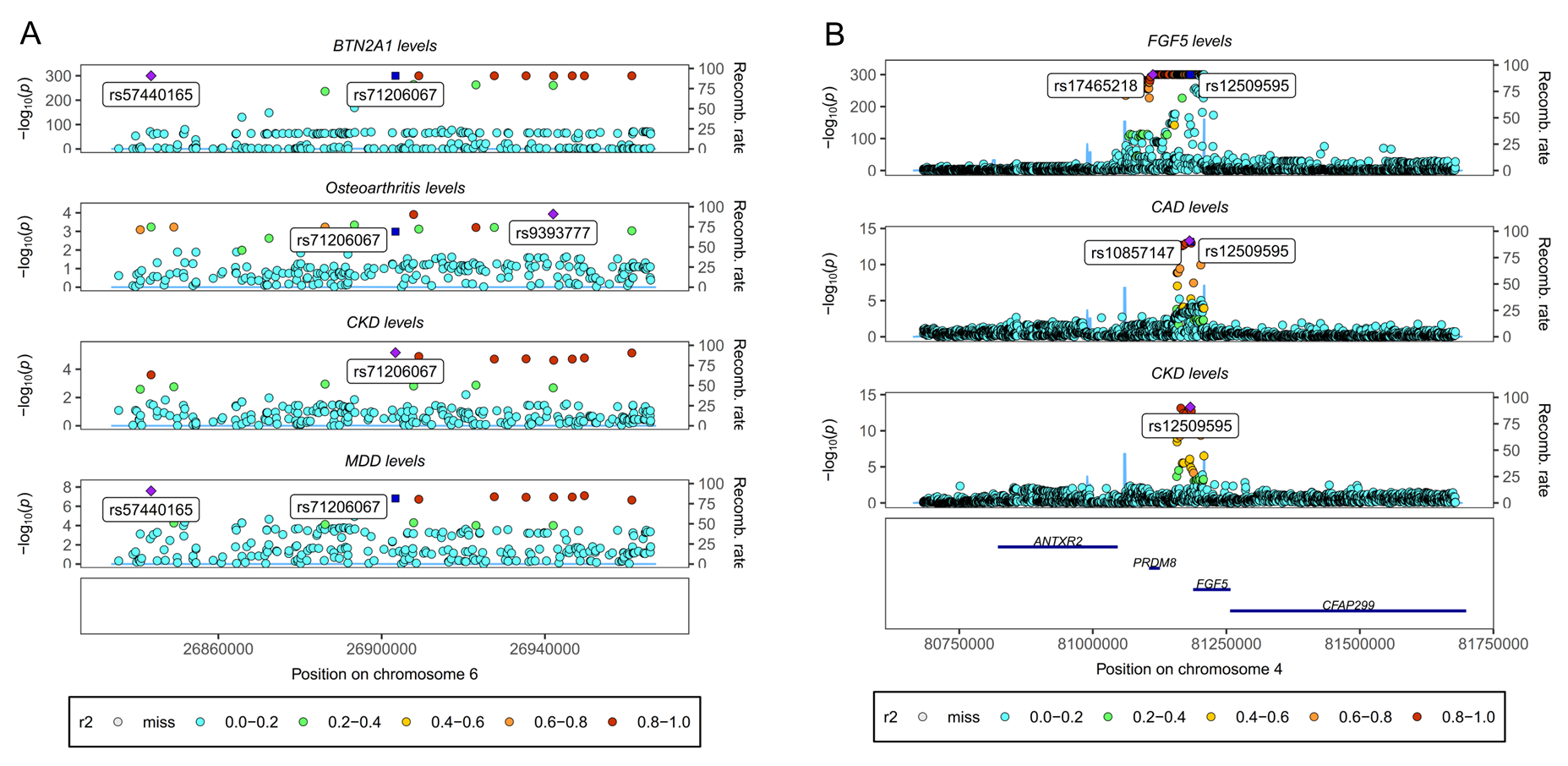

**Supplementary Figure 1**. Colocalization between pQTL of identified protein targets and multimorbidity GWAS with high or moderate support of colocalization evidence.

**(A)** Colocalization of BTN2A1 pQTL signal in Osteoarthritis, CKD, and MDD GWAS signal at the BTN2A1 locus. **(B)** Colocalization of FGF5 pQTL signal in CAD and CKD GWAS signal at the FGF5 locus.

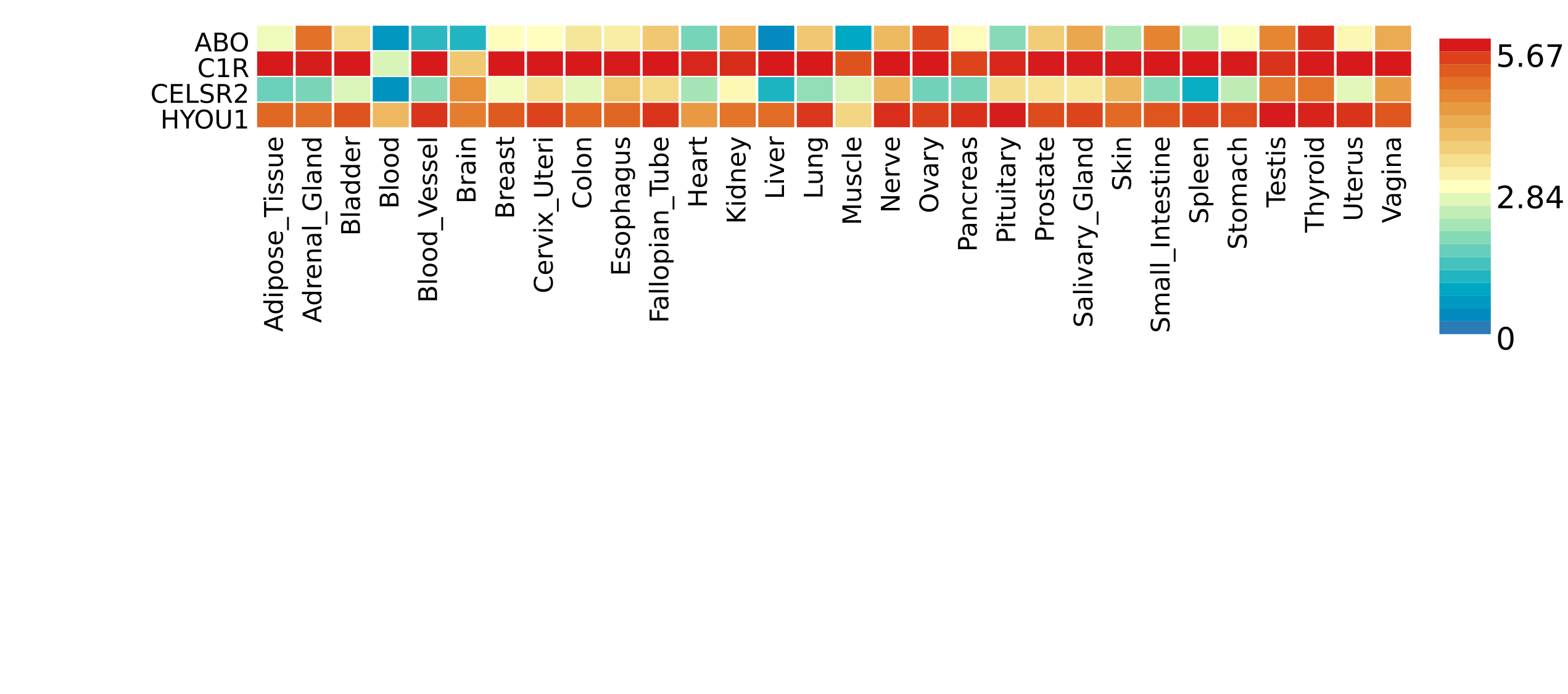

**Supplementary Figure 2**. Heatmap plot for tissue-specific expression of four identified *potential protein targets*. Gene expression levels were calculated by adding a pseudocount of 1 to the raw expression values, followed by log2 transformation. The average expression was then computed across 30 distinct tissue types.

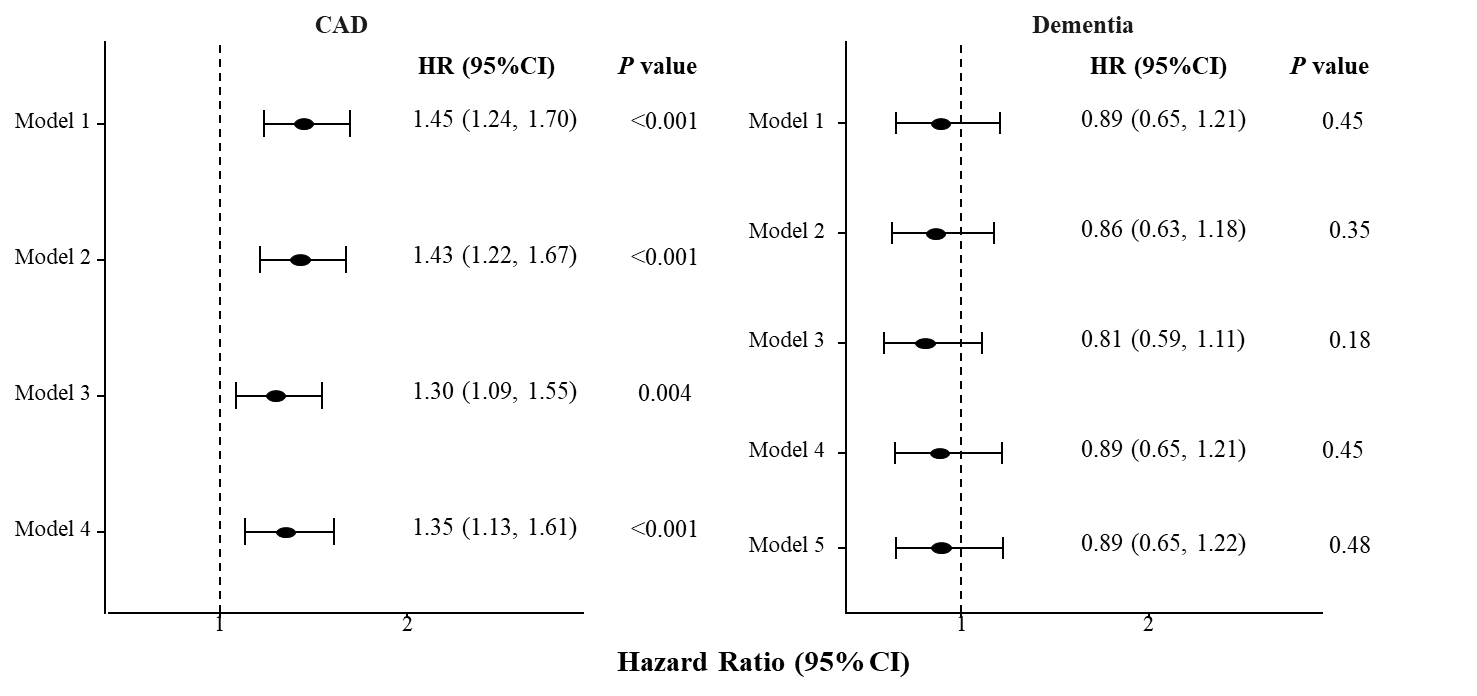

**Supplementary Figure 3. Associations of plasma C1R levels with incident CAD and dementia in the UK Biobank.**

During follow-up, 3,099 incident CAD events occurred over 586,216.2 person-years among 50,349 participants, and 1,392 incident dementia events occurred over 716,469.1 person-years among 52,958 participants. Model 1 was adjusted for age (continuous), sex (male or female), self-reported race (White, Asian or Asian British, Black or Black British, or other), and study center (England, Wales, or Scotland). Model 2 was further adjusted for educational attainment (college or university degree, intermediate qualifications, or low qualifications) and Townsend deprivation index (continuous). Model 3 was further adjusted for body mass index (<25, 25–30, or ≥30 kg/m²), drinking status (never, previous, or current), smoking status (never, former, or current), and physical activity (low, moderate, or high). Model 4 was further adjusted for hypertension, hyperlipidemia, and cancer (yes or no). Model 5 was further adjusted for cardiovascular disease (yes or no), social isolation (yes or no), sleep quality (good or poor), depressive symptoms (yes or no), family history of dementia (yes or no), and APOE ε4 carrier status (yes or no).

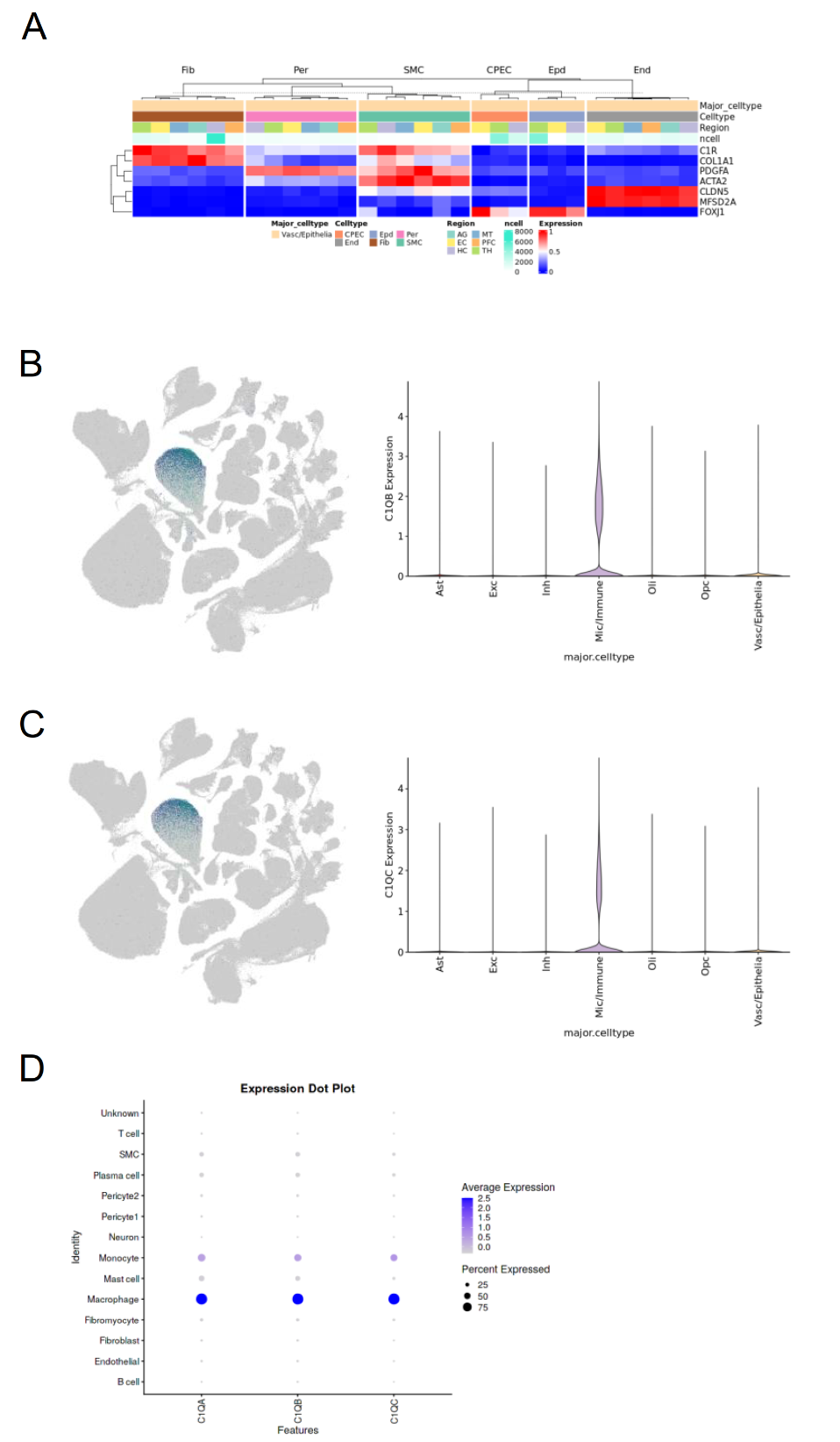

**Supplementary Figure 4.** Distribution of C1R and C1Q in dementia and cardiovascular disease

(A) Pseudobulk analysis showing enrichment of C1R, COL1A1, PDGFA, ACTA2, CLDN5, MFSD2A, and FOXJ1 gene expression across fibroblasts, pericytes, smooth muscle cells, choroid plexus epithelial cells, ependymal cells, and endothelial cells within the vascular/epithelial cell cluster of Alzheimer’s disease brain tissue.

(B, C) UMAP and violin plots showing the expression distribution of C1QB and C1QC in Alzheimer’s disease brain tissues.

(D) Dot plot illustrating the distribution of C1QA, C1QB, and C1QC expression in atherosclerotic vascular tissues.

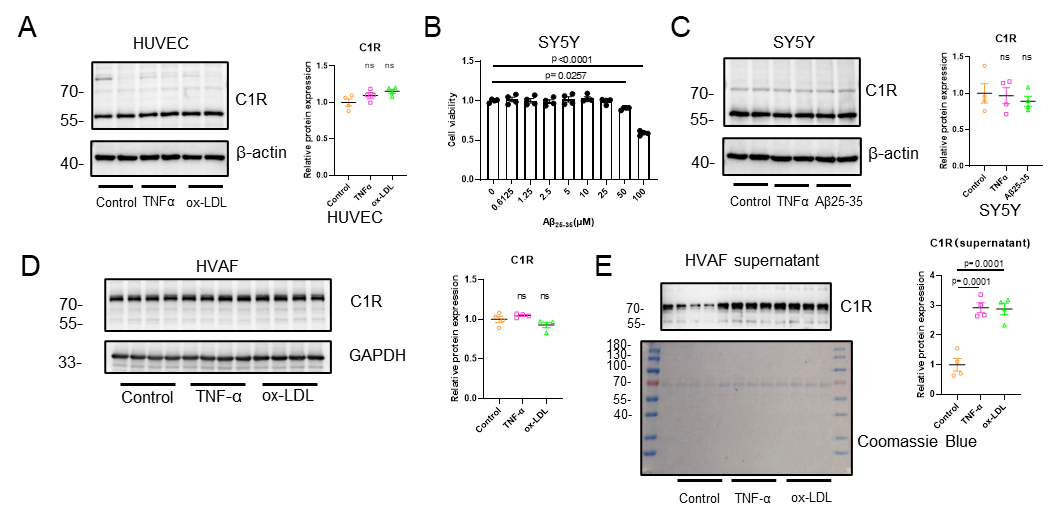

**Supplementary Figure 5**. C1R exacerbates vascular endothelial inflammation and neuronal injury

(A–D) Relative protein expression of C1R in HUVECs, SH-SY5Y, or HVAFs cells following stimulation with TNF-α and ox-LDL, or TNF-α and Aβ25–35 (n = 4 per group).

(E) Quantification of C1R protein levels in conditioned media from HVAFs following TNF-α and ox-LDL stimulation (n = 4 per group).
